# Closing Gaps in Evidence-Based Care for Acute Myocardial Infarction in northern Tanzania: Insights from a Prospective Study

**DOI:** 10.1101/2024.12.12.24318909

**Authors:** Olanrewaju Adisa, Erin E Brown, Francis M Sakita, Gloria Temu, Anzibert Rugakingira, Godfrey Lameck, Frida M Shayo, Blandina T Mmbaga, Nathan M Thielman, Gerald S Bloomfield, Janet P Bettger, Julian T Hertz

## Abstract

**Background:** The burden of Acute Myocardial Infarction (AMI) is growing in sub-Saharan Africa. In Tanzania, uptake of diagnostic testing and evidence-based therapy for AMI is suboptimal. We aimed to describe current gaps in evidence-based AMI care in a Tanzanian emergency department (ED) and estimate the potential benefit of closing key performance gaps.

**Methods:** Adults presenting with chest pain or dyspnea to the Kilimanjaro Christian Medical Centre (KCMC) ED were prospectively enrolled from February to September 2023 and their diagnostic tests and treatments were recorded. Thirty days following enrollment, a follow-up telephone survey was administered to assess mortality and medication use. Key performance metrics included the proportion of participants receiving both electrocardiography (ECG) and cardiac biomarker testing, as well as the proportion of participants with AMI receiving evidence-based therapies. To estimate the benefits of closing performance gaps, the the annualized number of participants not receiving each element evidence-based therapy was divided by published numbers needed to treat (NNTs) for each intervention. An exploratory analysis was conducted using performance metrics at KCMC and published national incidence data to estimate the potential benefits of closing performance gaps in AMI care at scale across Tanzania.

**Results:** Of 275 enrolled participants, 41 (14.9%) met criteria for AMI. Of participants, 91 (33.1%) received both ECG and cardiac biomarker testing. Of participants with AMI, 14 (34%) received aspirin, 11 (27%) received clopidogrel, and 2 (5%) received heparin. At 30-day follow-up, 25 (61%) participants with AMI were still alive, of whom 3 (12%) were taking dual antiplatelet therapy. Closing the diagnostic gap at KCMC could lead to the identification of an estimated 121 additional AMI cases annually. Aggregate estimated total benefit for closing gaps in uptake of aspirin, clopidogrel, heparin, and dual-antiplatelet therapy would result in approximately 6 deaths prevented and 33 serious cardiovascular events prevented at KCMC annually. Closing performance gaps at scale across Tanzania would result in approximately 69,206 additional AMI cases identified, 3,003 deaths prevented, and 11,405 serious cardiovascular events prevented annually.

**Conclusions:** Closing diagnostic and treatment gaps in AMI care in northern Tanzania presents an opportunity to increase case identification, save lives, and prevent cardiovascular events and rehospitalizations.

## Introduction

The rising burden of acute myocardial infraction (AMI) poses a public health threat in sub-Saharan Africa (SSA).^1^ In Tanzania, for example, the burden of AMI has escalated rapidly, making it the nation’s second leading cause of noncommunicable disease death behind stroke.^2^ As AMI becomes increasingly common, local health systems, which have historically prioritized communicable diseases, will need to adjust to address this rising tide of cardiovascular disease.

Low-cost, evidence-based interventions, such as routine electrocardiogram (ECG) and troponin testing, early aspirin administration, and dual anti-platelet therapy, reduce AMI morbidity and mortality.^3^ While the merits of evidence-based AMI care are well-established, implementation of such protocols in SSA remains sub-optimal.^3–8^ In northern Tanzania, for example, preliminary investigations conducted in a regional emergency department (ED) found that AMI is commonly misdiagnosed and inappropriately treated, resulting in high morbidity and mortality.^7^ A prospective study of AMI in the region in 2018 revealed that less than 3% of ED patients with possible AMI symptoms received ECG and troponin testing, approximately 90% of AMI cases were missed, only 21% of AMI patients received aspirin, and less than 5% received clopidogrel or heparin.^5, 7^ Low uptake of evidence-based care resulted in a 30-day mortality rate of 43% among AMI patients, eclipsing mortality rates published in any other recent study globally.^7, 8^ These findings, coupled with the projected growth in AMI burden, portend an escalation in AMI-associated mortality and morbidity in Tanzania if measures to promote evidence-based care are not implemented.

In response to these findings, an interdisciplinary team has been developing an intervention to improve AMI care at a referral hospital in northern Tanzania, Kilimanjaro Christian Medical Centre (KCMC).^9^ As part of our needs assessment, we conducted an observational study to describe current patterns of care for AMI at KCMC to identify gaps to target with our intervention. Because ample data is available regarding the benefits of evidenced-based AMI care, this gap analysis allowed us to estimate the potential benefits of a quality improvement (QI) program.

## Methods

### Setting

This study was conducted in the ED at KCMC in the northern Tanzania. KCMC serves a catchment area with nearly 15 million individuals and was the site for our team’s preliminary studies of AMI burden and outcomes.^1, 5, 7, 8^ The KCMC ED has 24-hour access to basic AMI diagnostic technology such as ECG, laboratory-based and point-of-care troponin testing, and echocardiography. However, KCMC did not have formally trained cardiologists nor capacity for percutaneous coronary intervention (PCI) at the time of this study. The nearest facility with PCI capability is approximately 10 hours from KCMC by motor vehicle. The KCMC ED serves all patients presenting to the hospital for urgent and emergent care, without referral.

### Participant Selection

Inclusion criteria and enrollment procedures were identical to prior studies of AMI at KCMC.^1, 5, 7, 8^ All patients aged ≥18 years presenting to the ED were screened, and those with with chest pain or shortness of breath as a primary or secondary complaint were offered enrollment. We excluded patients with any of the following: (1) inability to provide informed consent, (2) traumatic chest pain, or (3) self-reported fever. This study was conducted between February 1^st^ and September 1^st^, 2023, with enrollment conducted seven days per week from 8AM until 10PM.

### Study procedures

After obtained informed consent, research staff used a standardized data collection form to record patient sociodemographic information, past medical history, and history of present illness. The decision to obtain an ECG and order troponin testing for participants was made by the treating ED physician based on clinical discretion. If the ED clinical team did obtain an ECG on any participant, a digital photograph of the ECG was obtained by the research team. The research team also collected the results of any laboratory testing obtained by the ED team, including troponin and creatinine levels, directly from the electronic medical record.

Research assistants directly observed, completed electronic medical record chart reviews for, and documented all treatment and diagnostic testing that participants received while in the ED, including administration of aspirin, clopidogrel, and heparin. For participants who were hospitalized, the final hospital discharge diagnoses were also collected from the electronic medical record. Thirty days following enrollment, participants were contacted via telephone to assess vital status, symptom progression, and medication use. If the team was unable to contact participants via telephone, a research assistant visited the participant’s home to administer the follow-up questionnaire in person.

### ECG interpretation

All ECGs were shared immediately with the clinical team. For study purposes, two independent physicians from the research team, with residency training in emergency medicine, interpreted all ECGs. Physician adjudicators determined the presence of ST-elevation myocardial infarction (STEMI) using the Fourth Universal Definition of Myocardial Infarction guidelines and the modified Sgarbossa criteria.^10, 11^ Agreement among adjudicators for presence of STEMI was 97.4%. In instances of disagreement, a third physician adjudicator was used as the tiebreaker.

### Study definitions

The criteria for defining AMI were derived from the Fourth Universal Definition of Myocardial Infarction,^10^ as summarized in Table 1. AMI was defined by a final hospital discharge diagnosis of AMI, the presence STEMI on ECG, non-STEMI as indicated by an abnormally elevated troponin level paired with an abnormal three-hour delta troponin, or a single abnormally elevated troponin value in the absence of advanced renal dysfunction (glomerular filtration rate <15 ml/min/1.73m^2^) if the clinical team did not obtain serial troponins.

**Table 1.**
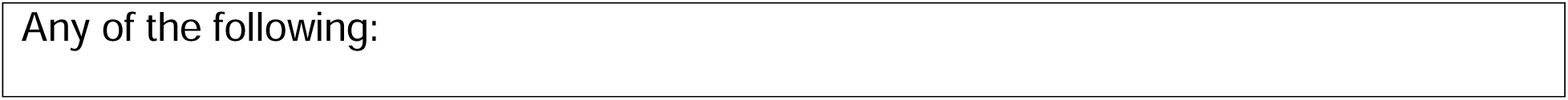
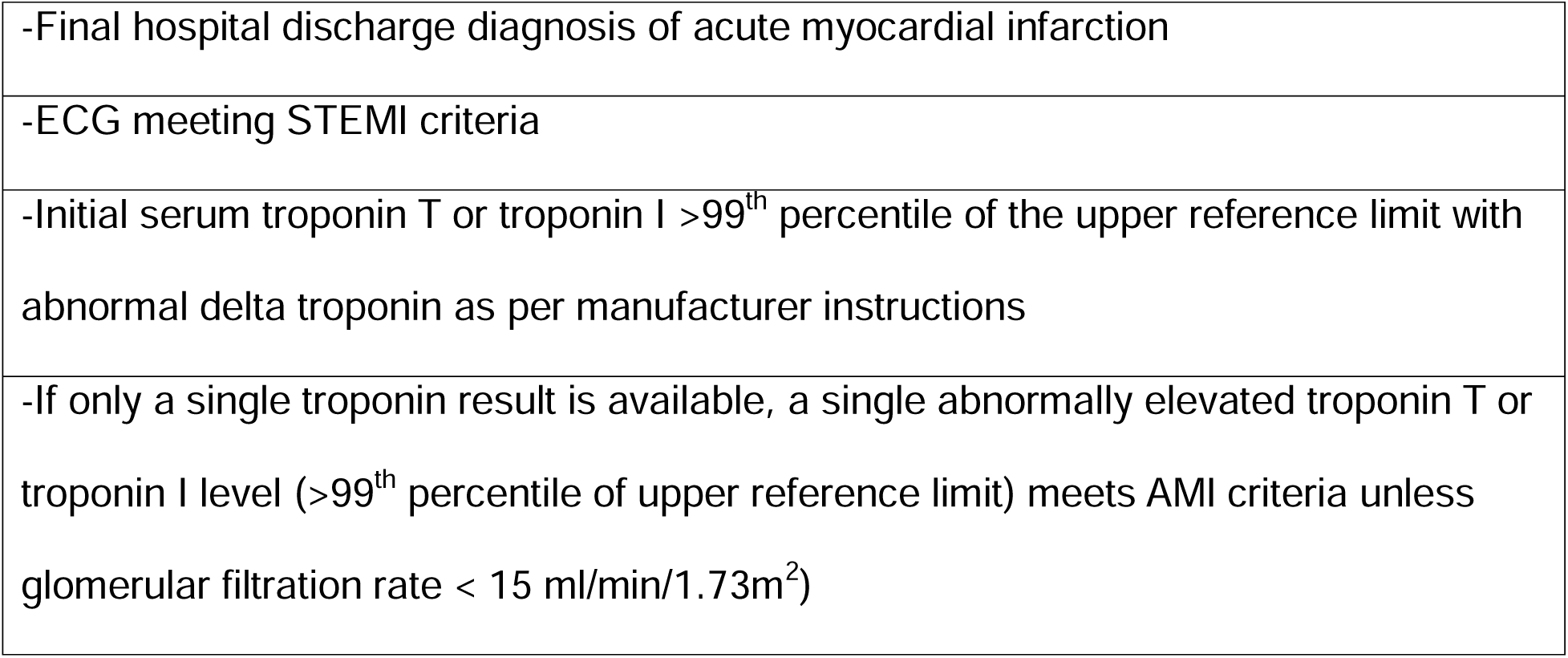
Study definition for acute myocardial infarction (AMI)

|  |
| --- |
| -Final hospital discharge diagnosis of acute myocardial infarction |
| -ECG meeting STEMI criteria |
| -Initial serum troponin T or troponin I >99<sup>th</sup> percentile of the upper reference limit with abnormal delta troponin as per manufacturer instructions |
| -If only a single troponin result is available, a single abnormally elevated troponin T or troponin I level (>99<sup>th</sup> percentile of upper reference limit) meets AMI criteria unless glomerular filtration rate < 15 ml/min/1.73m<sup>2</sup>) |

Baseline participant diabetes was defined by any of the following: self-reported history of diabetes, fasting glucose ≥ 126 mg/dl, or random glucose ≥ 200 mg/dl. Other co-morbidities, including hypertension, HIV infection, and prior MI were defined by participant self-report. Five-year risk of cardiovascular event was determined by the Harvard National Health And Nutrition Examination Survey formula, based on participant age, gender, smoking status, diabetes, and systolic blood pressure.^12^ Dual anti-platelet therapy was defined as use of aspirin and any other antiplatelet agent, such as clopidogrel. Serious cardiovascular event was defined as recurrent myocardial infarction, stroke, or hospitalization due to heart failure.^13–19^

### Statistical analyses

Nine key performance metrics were calculated: the proportion of participants who received ECG testing, the proportion of participants who received cardiac biomarker testing (defined as receiving at least one troponin test), the proportion of participants who received both ECG and cardiac biomarker testing, the proportion of participants meeting AMI criteria who received aspirin in the ED, the proportion of participants with AMI who received clopidogrel in the ED, the proportion of participants with AMI who received heparin in the ED, the proportion of participants with AMI surviving to 30 days, the proportion of participants with AMI who were taking aspirin 30 days following enrollment, and the proportion of participants taking dual antiplatelet therapy 30 days following enrollment. Estimated glomerular filtration rate was calculated from measured serum creatinine using the race-neutral 2021 CKD-EPI equation.^20^ All statistical analyses were performed in the R Suite.

To estimate the potential benefits of closing performance gaps for the nine key metrics, additional calculations were made. First, the number of additional AMI cases that could be identified in a year by increasing testing uptake was calculated by multiplying the number of participants who did not receive complete AMI testing (ECG and at least one troponin assay) by 22.3% (the proportion of KCMC ED patients with chest pain or dyspnea who met AMI criteria when universal testing was performed).^7^ This number was further multiplied by 1.71 (to account for overnight hours when enrollment was not performed) and 1.72 (to account for the seven-month enrollment period) to obtain an annualized figure. The resulting number was assumed to represent the total number of AMI cases that were missed by usual care annually, which also represents the total number of additional AMI cases that could be identified by increasing testing uptake to 100% of patients with potential AMI symptoms.

To estimate the potential benefits of increasing uptake of aspirin, clopidogrel, and heparin, a literature search was performed to determine the number needed to treat (NNT) for each of these evidence-based therapies.^13–18^ The NNTs obtained from the literature are presented in Table 2. The annualized number of AMI patients not receiving each evidence-based therapy was adjusted to account for overnight hours and the seven-month enrollment period, as described above. The resulting number was then added to the total number of annual missed AMI cases (described above), who were assumed to not have received the evidence-based therapy. The sum of these numbers represented the estimated total number of patients with AMI who did not receive the evidence-based therapy each year. This total number was divided by the NNT to estimate the annualized potential benefit of achieving 100% uptake of each evidence-based therapy. Detailed explanations of these calculations are provided in the supplementary material (See Supplement).

**Table 2.** Number Needed to Treat (NNT) for Evidence-Based Therapies for Acute Myocardial Infarction (AMI)

| Intervention | Benefit | NNT |
| --- | --- | --- |
| Aspirin for AMI | Prevented death<sup>13</sup> | 42 |
|  | Serious cardiovascular event<br>(recurrent MI) prevented<sup>19</sup> | 29 |
| Aspirin for secondary<br>prevention following<br>AMI | Serious cardiovascular event<br>prevented<sup>15</sup> | 26 |
|  | Prevented death<sup>14</sup> | 333 |
| Heparin for AMI | Prevented recurrent MI<sup>16</sup> | 33 |
| Clopidogrel for AMI | Serious cardiovascular event<br>prevented<sup>17, 18</sup> | 23 |
| Clopidogrel in addition<br>to aspirin for<br>secondary prevention<br>following AMI (Dual<br>Antiplatelet therapy) | Prevented death<sup>18</sup> | 250 |
|  | Serious cardiovascular event<br>prevented<sup>18</sup> | 43 |

In addition, we conducted an exploratory supplemental analysis to estimate the potential benefits of closing performance gaps in AMI care at scale across Tanzania. To do so, we used published estimates of the national population^21^ and the annual incidence of AMI in Tanzania^1^ to estimate the annual number of AMI cases in Tanzania. We assumed that care observed at KCMC is representative of typical AMI care across Tanzania. We then calculated the proportion of AMI patients at KCMC receiving each evidence-based care element (complete AMI testing, aspirin, clopidogrel, heparin), and applied these proportions to the annual number of AMI cases in Tanzania to estimate the total number of AMI patients not receiving each therapy annually. Finally, we used published NNTs to estimate the potential benefit of closing these performance gaps across the country, using the same method described above. Detailed explanations of these calculations are provided in the supplementary material (See Supplement).

### Data availability

The data underlying this article will be shared on reasonable request to the corresponding author.

## Results

During the study period, a total of 2725 adult ED patients were screened and 276 (10.1%) were eligible for study inclusion (Figure 1). Of these, all but one patient consented to participate in the study and were enrolled. Table 3 presents the characteristics of the study participants. The median (IQR) age of participants was 63 (46.5, 76.5) years, and more than half (n=160) of study participants were female (58.4%). The most common self-reported medical comorbidities among participants were hypertension (n=165, 60.0%) and diabetes (n=62, 22.5%). Slightly over one-third of all patients had a five-year cardiovascular event risk of greater than 30% (n=96, 34.9%). Approximately one third of all participants had a prior MI or stroke (n = 92, 33.5%).

**Figure 1:**
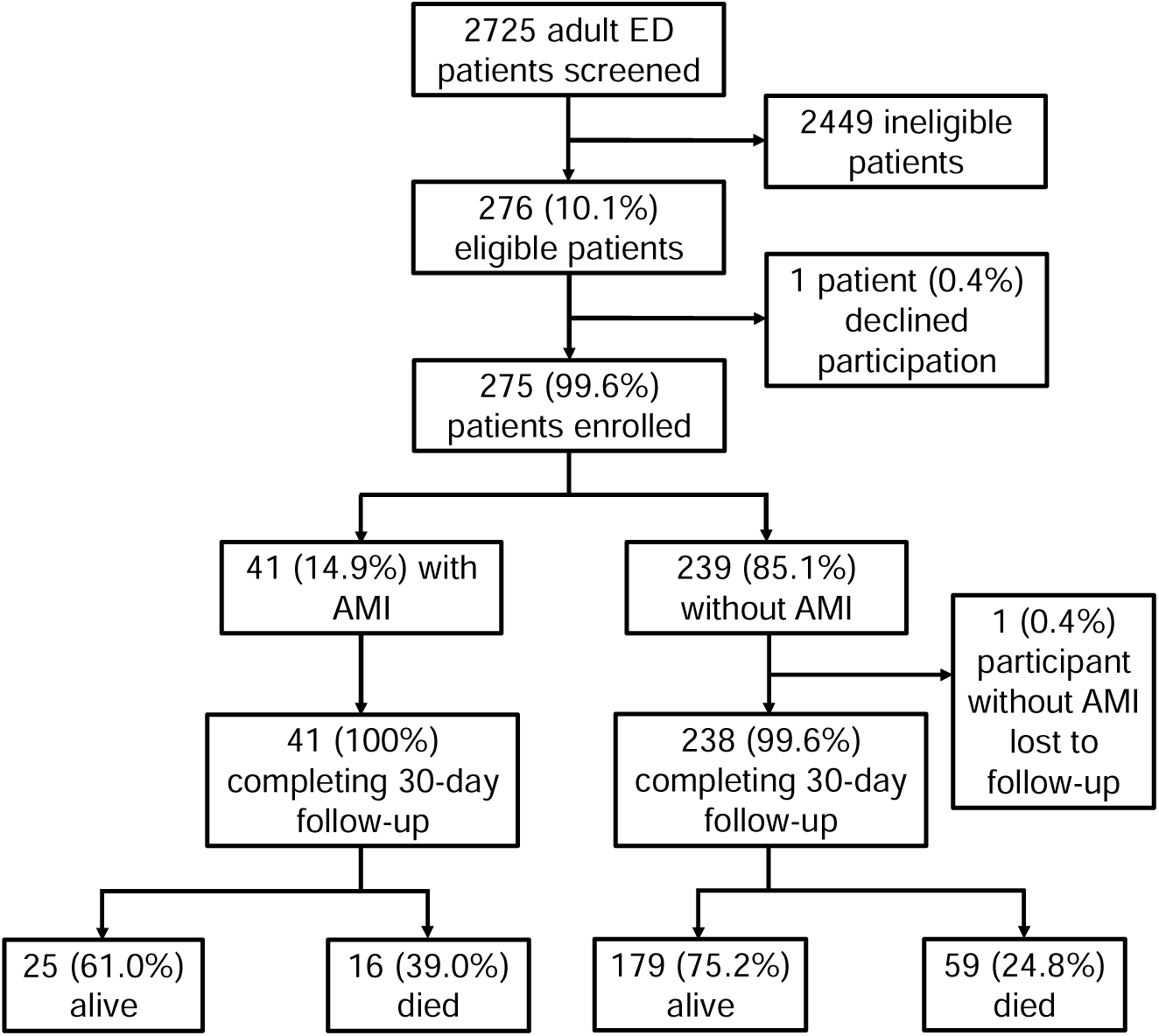
Flow diagram of study participants

**Table 3.**
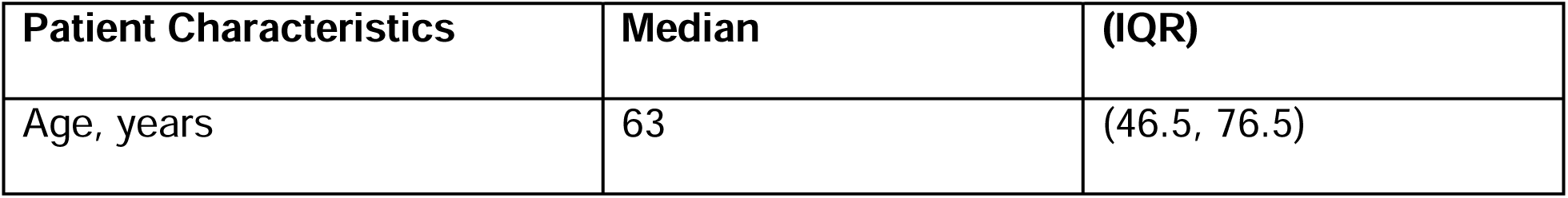
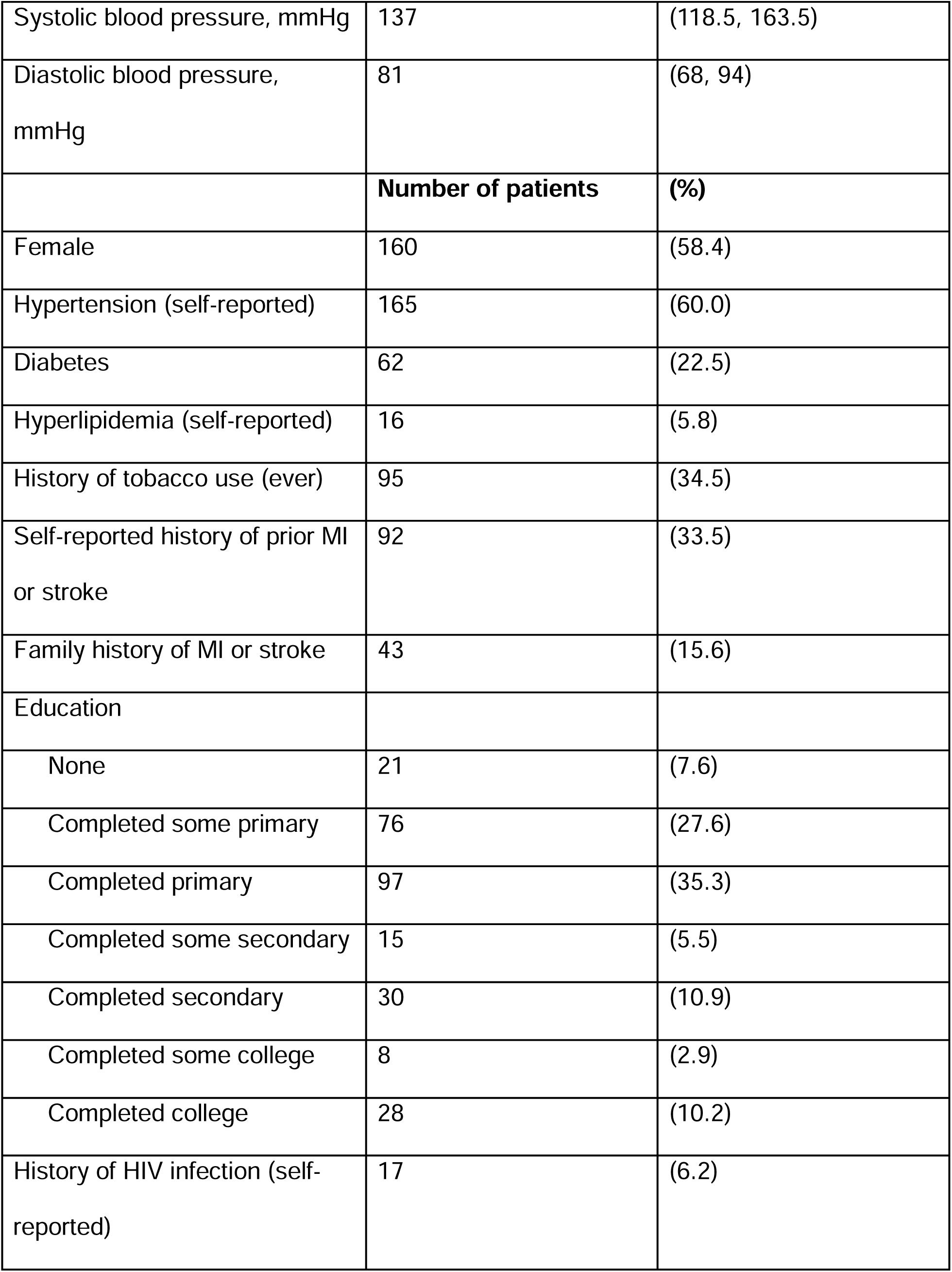
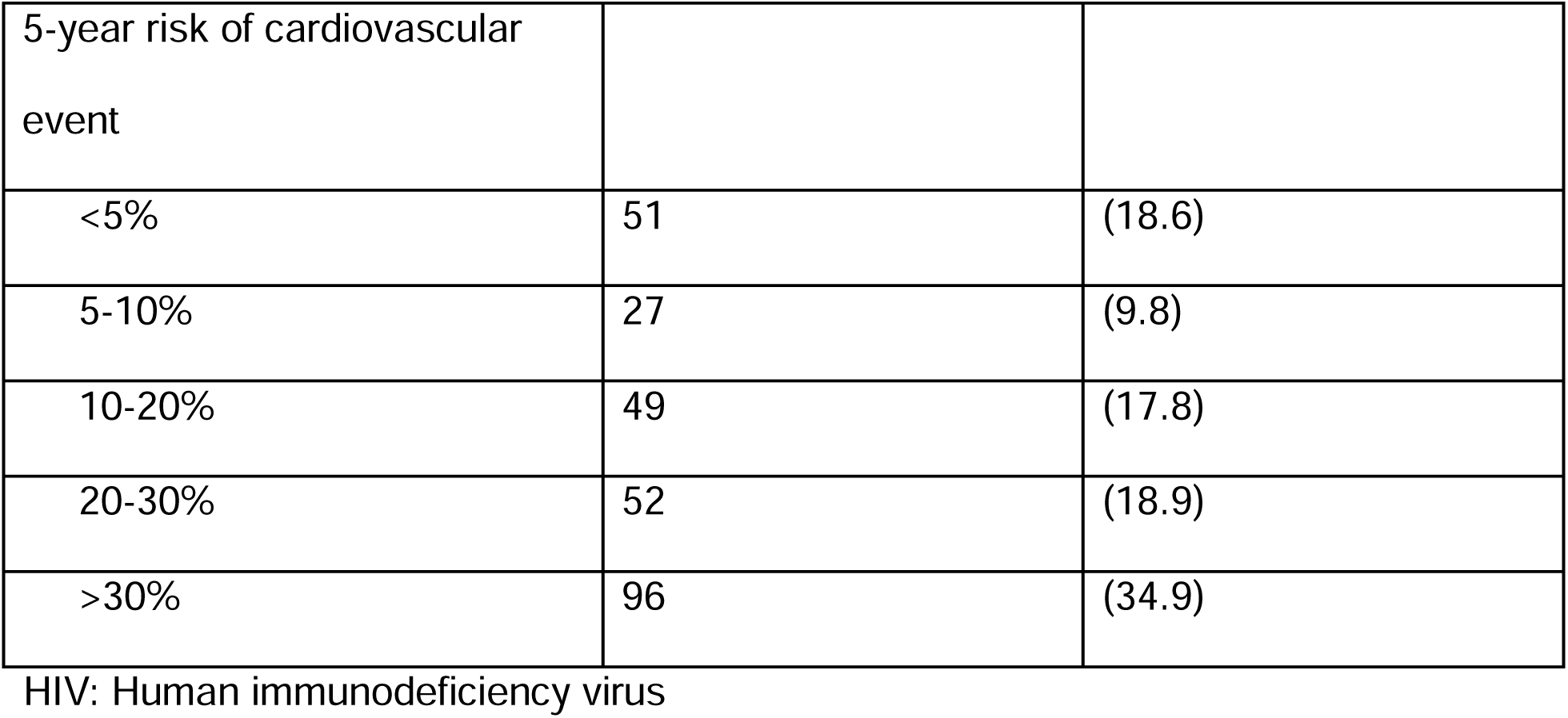
Characteristics and cardiovascular risk factors among emergency department patients with chest pain or shortness of breath, northern Tanzania, 2023 (N=275)

| Patient Characteristics | Median | (IQR) |
| --- | --- | --- |
| Age, years | 63 | (46.5, 76.5) |
| Systolic blood pressure, mmHg | 137 | (118.5, 163.5) |
| Diastolic blood pressure, mmHg | 81 | (68, 94) |
|  | **Number of patients** | **(%)** |
| Female | 160 | (58.4) |
| Hypertension (self-reported) | 165 | (60.0) |
| Diabetes | 62 | (22.5) |
| Hyperlipidemia (self-reported) | 16 | (5.8) |
| History of tobacco use (ever) | 95 | (34.5) |
| Self-reported history of prior MI or stroke | 92 | (33.5) |
| Family history of MI or stroke | 43 | (15.6) |
| Education |  |  |
| None | 21 | (7.6) |
| Completed some primary | 76 | (27.6) |
| Completed primary | 97 | (35.3) |
| Completed some secondary | 15 | (5.5) |
| Completed secondary | 30 | (10.9) |
| Completed some college | 8 | (2.9) |
| Completed college | 28 | (10.2) |
| History of HIV infection (self-reported) | 17 | (6.2) |
| 5-year risk of cardiovascular event |  |  |
| <5% | 51 | (18.6) |
| 5-10% | 27 | (9.8) |
| 10-20% | 49 | (17.8) |
| 20-30% | 52 | (18.9) |
| >30% | 96 | (34.9) |

HIV: Human immunodeficiency virus

Of participants, 41 (14.9%) met the study definition for AMI. One patient without AMI was lost to follow up, but 30-day follow-up was achieved for all other participants (99.6% overall follow-up rate).

Table 4 presents the summary of key performance metrics. Of the 275 participants with chest pain or dyspnea, 152 (55.3%) received ECG testing, 114 (41.4%) received cardiac biomarker testing, and 91 (33.1%) received both ECG and cardiac biomarker testing. Of the 41 participants with AMI, 14 (34.1%) received aspirin in the ED, 11 (26.8%) received clopidogrel, and 2 (4.9%) received heparin. Of participants with AMI, 25 (61%) survived to 30-day follow up. Of the 25 surviving AMI participants, 4 (16.0%) were taking daily aspirin thirty days following enrollment while 3 (12.0%) were taking dual antiplatelet therapy.

**Table 4.** Summary of Key Performance Metrics.

| **Key Performance Metrics** | (n/N) | (%) |
| --- | --- | --- |
| Received ECG testing | 152/275 | 55.3 |
| Received cardiac biomarker testing | 114/275 | 41.4 |
| Received ECG testing and cardiac biomarker testing | 91/275 | 33.1 |
| Received aspirin in the ED* | 14/41 | 34.1 |
| Received clopidogrel in the ED* | 11/41 | 26.8 |
| Received heparin in the ED* | 2/41 | 4.9 |
| Participants surviving to 30 days* | 25/41 | 61.0 |
| Participants taking aspirin 30 days following enrollment* | 4/25 | 16.0 |
| Participants taking dual antiplatelet therapy 30 days following enrollment* | 3/25 | 12.0 |

\* among participants diagnosed with AMI
\*\* among participants diagnosed with AMI who survived past 30 days

The annualized potential benefits of closing the performance gaps for each key evidence-based care metric are summarized in Table 5. An estimated total of 121 additional AMI cases could be identified by increasing testing uptake to 100% of patients with AMI symptoms. Aggregate estimated total benefit for closing gaps in use of aspirin, clopidogrel, heparin, and dual-antiplatelet therapy would result in approximately 6 deaths prevented and 33 serious cardiovascular events prevented annually.

**Table 5.**
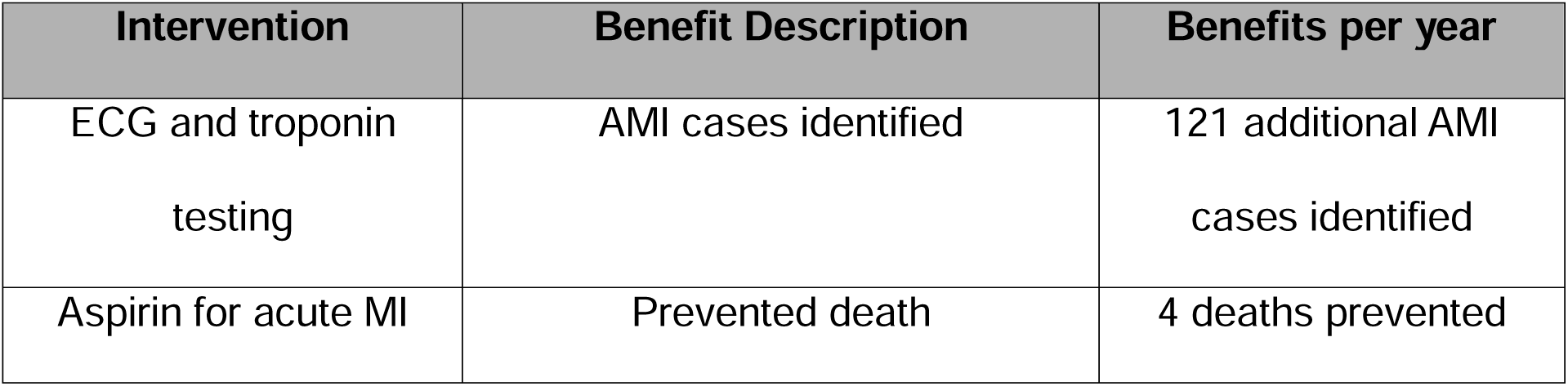
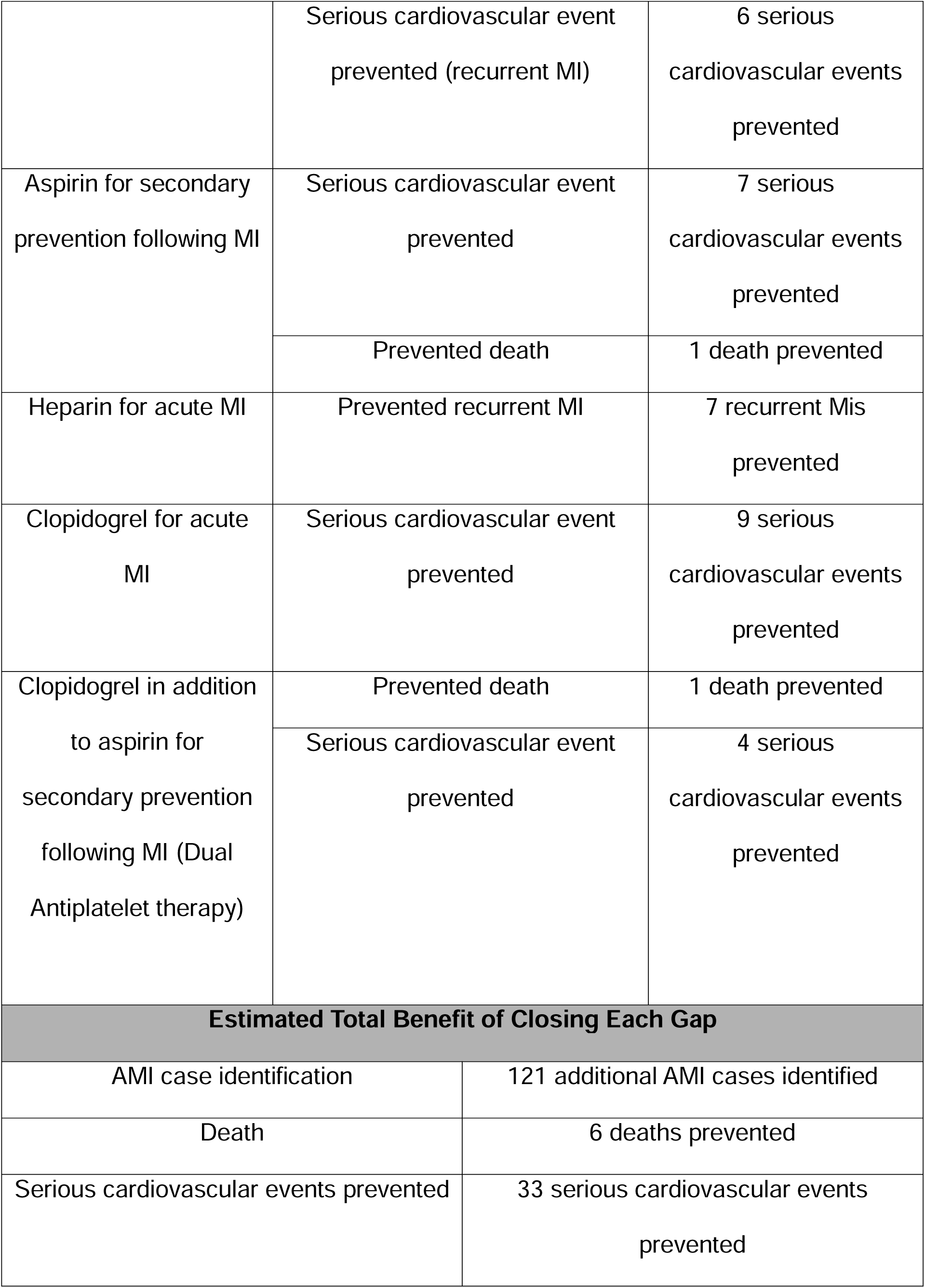
Annualized potential benefit of closing gaps in evidence-based therapies at KCMC.

The annualized potential benefits of closing the performance gaps for each key evidence-based metric at scale across Tanzania can be found in Supplementary Table 3 (see Supplement). Closing performance gaps in AMI testing across Tanzania would result in identification of an estimated additional 69,206 AMI cases. Aggregate estimated total benefits for closing gaps in use of aspirin, clopidogrel, heparin, and dual-antiplatelet therapy would result in approximately 3,003 deaths prevented and 11,405 serious cardiovascular events prevented across Tanzania annually.

## Discussion

### Statement of principal findings

This study is part of an interdisciplinary quality improvement initiative to improve AMI care at KCMC. As part of our needs assessment,^9^ we analyzed current quality gaps and estimated the potential benefit of closing such gaps. In doing so, we highlighted the urgent need for a quality improvement intervention for AMI care in our setting, which has the potential to increase AMI case identification, prevent serious cardiovascular events, and save lives.

### Interpretation within the context of the wider literature

Our results revealed substantial gaps in testing of patients presenting with potential AMI symptoms. Because AMI can present with subtle symptoms that overlap with many other conditions, international guidelines recommend broad AMI screening criteria in the ED to maximize case identification.^3, 4, 22^ Almost half of patients presenting with potential AMI symptoms in our study did not undergo ECG testing, and only one-third of patients received both ECG and cardiac biomarker testing, as recommended by international guidelines.^22^ Closing this diagnostic gap would result in the identification of approximately 121 additional AMI cases per year at our center—a substantial opportunity to increase case identification and initiate appropriate therapies. Further, our supplemental analysis suggests that closing the diagnostic gap at scale across Tanzania could identify an additional 69,206 cases per year. Our findings add to a growing body of evidence suggesting that under-diagnosis of AMI in SSA is common, resulting in under-estimation of disease burden.^5, 7, 23^ Such underestimation further reinforces the misperception that AMI is rare in SSA, which in turn may further bias local clinicians against AMI testing.

Our analysis revealed opportunities for improvement in use of evidence-based AMI therapies at KCMC. Consistent with prior studies in Tanzania,^1, 5, 7, 8^ less than one-third of AMI patients received aspirin in the ED and even fewer received clopidogrel or heparin. The potential impact of closing these performance gaps is substantial, with our supplemental analysis estimating over 3000 potential lives saved and 11,000 serious cardiovascular events prevented annually across the country. Studies suggesting that other communities in Tanzania have both higher burden of AMI risk factors and more limited access to AMI care.^24, 25^ Given these variations, our national at-scale estimates for lives saved and serious cardiovascular events prevented should be considered conservative, suggesting the possibility of even greater benefits from closing evidence-based treatment gaps across the country. Other studies in Ghana, Kenya, and Ethiopia identified similar gaps in guideline-recommended treatment,^6, 26, 27^ suggesting that there are likely widespread opportunities to improve AMI care and reduce associated morbidity and mortality across SSA.

### Implications for policy, practice, and research

Many factors may contribute to the observed performance gaps in AMI diagnosis and treatment. Complexity and cost of AMI care, hospital culture, lack of resources, inadequate knowledge about AMI among providers and patients, need for quality improvement leaders, poor doctor-patient communication, and inefficient care systems are key barriers to evidence-based AMI care identified in previous studies in Tanzania.^28, 29^ Given that providers in Tanzania report strongly positive attitudes towards quality improvements for AMI care,^30^ provider training may be one potential avenue for improved care and outcomers. Careful consideration of other factors leading to poor uptake of evidence-based AMI care can inform a multifaceted approach to improve systems for early identification and treatment of patients presenting with AMI symptoms in Tanzania.^9, 29^

### Strengths and limitations

We acknowledge several limitations of our analysis. Although we used NNT estimates from large randomized controlled trials, these trials were conducted in high-income settings with limited participant diversity.^13, 14, 16, 17, 19^ Although the benefits of evidence-based therapies like aspirin are well-established, the exact NNT for each therapy in our setting is unknown. Secondly, this study was conducted at a single ED in northern Tanzania, which limits the generalizability of our findings to other settings. Thirdly, we applied multipliers to account for times when enrollment was not conducted, assuming that there was no seasonal or hourly variation in frequency of AMI presentation. If there was variability in AMI presentation at KCMC by time of day or time of year, this may have resulted in imprecise estimation of performance gaps. Finally, to estimate the potential benefit of closing care gaps across Tanzania, we assumed that the care observed at KCMC was representative of AMI care across Tanzania. To our knowledge, there are no published studies reporting usual AMI care at other hospitals in Tanzania, and so the accuracy of this assumption is unknown. As many hospitals in Tanzania lack reliable access to ECGs and cardiac biomarker testing,^28, 29^ uptake of evidence-based care may be lower at other facilities, which would have resulted in an under-estimation of the benefit of closing performance gaps at scale across Tanzania.

## Conclusions

In conclusion, our performance gap analysis suggests the need for continued quality improvement initiatives to enhance AMI care in northern Tanzania. Our analysis highlighted the potential to significantly increase case identification, initiate appropriate therapies, and prevent cardiovascular events. The results of this study were used to inform the development of a quality improvement intervention^9^ which is currently being piloted at KCMC. Future studies will analyze the impact of this intervention on care processes and AMI outcomes.

## Data Availability

All data produced in the present study are available upon reasonable request to the authors

